# Circulating Low Density Neutrophils Are Associated with Resistance to First-Line Anti-PD1/PDL1 Immunotherapy in Non-Small Cell Lung Cancer

**DOI:** 10.1101/2022.04.27.22273598

**Authors:** H Arasanz, A Bocanegra, I Morilla, J Fernández-Irigoyen, M Martínez-Aguillo, L Teijeira, M Garnica, E Blanco, L Chocarro, K Ausin, M Zuazo, G Fernández-Hinojal, M Echaide, L Fernández-Rubio, S Piñeiro-Hermida, P Ramos, L Mezquita, D Escors, R Vera, G Kochan

**Affiliations:** Oncoimmunology Group, Navarrabiomed. Instituto de Investigación Sanitaria de Navarra (IdiSNA). Irunlarrea st., 3. 31008, Pamplona, Spain; Medical Oncology Department, Hospital Universitario de Navarra. Instituto de Investigación Sanitaria de Navarra (IdiSNA). Pamplona, Spain; Oncobiona Group, Navarrabiomed. Instituto de Investigación Sanitaria de Navarra (IdiSNA). Pamplona, Spain; Clinical Neuroproteomics Unit, Proteomics Platform, Proteored-ISCIII. Navarrabiomed. Instituto de Investigación Sanitaria de Navarra (IdiSNA). Pamplona, Spain; Gene Therapy and Regulation of Gene Expression, Centro de Investigación Médica Aplicada (CIMA). Instituto de Investigación Sanitaria de Navarra (IdiSNA). Pamplona, Spain; Medical Oncology Department, Hospital Clínico San Carlos. Madrid, Spain; Medical Oncology Department, Hospital Clínic i Provincial de Barcelona, IDIBAPS. Barcelona, Spain; Medical Oncology Department, Laboratory of Translational Genomics and Targeted Therapies in Solid Tumors, IDIBAPS. Barcelona, Spain

**Keywords:** Lung cancer, NSCLC, immunotherapy, neutrophils, LDN, biomarkers, PD-1, immunecheckpoint inhibitors

## Abstract

**Background:** Single-agent immunotherapy has been widely accepted as frontline treatment for advanced non-small cell lung cancer (NSCLC) with high tumor PD-L1 expression, however most patients do not respond and the mechanisms of resistance are not well known. Several works have highlighted the immunosuppressive activities of myeloid subpopulations, including low-density neutrophils (LDNs), although the context in which these cells play their role is not well defined.

**Methods:** We prospectively monitored LDNs in peripheral blood from patients with NSCLC treated with anti-PD-1 immune checkpoint inhibitors (ICIs) as frontline therapy, and correlated values with outcomes. We have monitored LDNs in a cohort of patients treated with anti-PD1 immunotherapy combined with chemotherapy (CT+IT). We performed ex vivo experiments, including comparative proteomics, to explore the underlying mechanisms.

**Results:** Elevated baseline LDNs predict primary resistance to ICI monotherapy in patients with NSCLC, with a response rate of 0% when levels are higher than 7.09%. Baseline LDNs are not associated with response to CT+IT. Quantitative proteomics of the plasma revealed an enrichment of the HGF/c-MET pathway in patients with high circulating LDNs, indicating a possible regulatory mechanism of this phenomenon.

**Conclusions:** Circulating LDNs mediate resistance in NSCLC receiving ICI as frontline therapy through humoral immunosuppression. A depletion of this population with CT+IT might overcome resistance, suggesting that patients with high PD-L1 tumor expression and high LDNs might benefit from this combination. The activation of the HGF/c-MET pathway in patients with elevated LDNs supports potential drug combinations targeting this pathway.

**TRANSLATIONAL RELEVANCE:** Immunotherapy has positioned as frontline therapy for advanced non-small cell lung cancer (NSCLC), alone when PD-L1 tumor expression is high, or combined with chemotherapy otherwise. However, 50% of the patients do not respond to the treatment and the mechanisms of resistance are not well defined. Moreover, it is not clear whether chemo-immunotherapy (CT+IT) could be advantageous in some patients with high PD-L1 tumor expression.

We have found that baseline circulating low-density neutrophils (LDN) identify a subset of patients that are intrinsically refractory to immunotherapy. Interestingly, responses can be achieved with CT+IT, detecting a progressive depletion of LDN in those patients. Besides the potential role as predictive biomarker, through ex vivo experiments and quantitative proteomics we observed that resistance was mediated by soluble molecules related with the HGF/c-MET pathway. Our findings establish circulating myeloid cells as one of the main mediators of resistance to immunotherapy in NSCLC, and gives a rationale for potential drug combinations that might improve the outcomes.

## INTRODUCTION

Immune checkpoint inhibitors (ICI) have improved the outcomes of patients diagnosed with advanced non-small cell lung cancer (NSCLC). Anti-PD1/PD-L1 antibodies have become the standard frontline treatment, as a single agent when PD-L1 tumor expression is high or combined with chemotherapy otherwise [1-8]. Unfortunately, the majority of the patients do not respond to treatment, no accurate predictive biomarkers able to identify those who will obtain a greater benefit have been discovered yet, and the mechanisms of resistance are not well understood [9-10]. Moreover, some patients with high PD-L1 expression who do not respond to single-agent ICI might benefit from chemoimmunotherapy combination.

Over recent years, the relevance of neutrophils in the immune response against cancer has been highlighted, although the understanding of their role is far from complete. Pro-tumor and anti-tumor activity have been described depending on the tissue and context [11-12]. A population of circulating neutrophils with immunosuppressive properties known as low-density neutrophils (LDNs) has been detected in cancer patients [13-15], although its association with response to ICI has not yet been defined.

We have used flow cytometry to prospectively monitor LDNs levels in fresh blood samples from a cohort of patients with advanced NSCLC treated with ICI monotherapy, and evaluated the association with response and disease control. These findings were compared with a cohort of patients treated with chemoimmunotherapy combination. Lastly, we explored ex vivo the mechanisms of resistance using co-cultures, and compared the plasma of the different cohorts of patients using quantitative proteomics.

## METHODS

### Study design and patient enrolment

The study was approved by the Ethics Committee of the University Hospital of Navarre. Informed consent was obtained from all subjects and all experiments were performed according to the principles stablished in the Declaration of Helsinki and the Department of Health and Human Services Belmont Report. Samples were collected by the Blood and Tissue Bank of Navarre, Health Department of Navarre, Spain. Thirty-one patients diagnosed of NSCLC and PD-L1 tumor expression ≥ 50% were recruited at the University Hospital of Navarre. All received anti-PD1 immunotherapy (pembrolizumab) as frontline therapy according to current indications. Twenty patients diagnosed of NSCLC and PD-L1 0-49% were recruited at University Hospital of Navarre. They were treated with platinum-based chemotherapy combined with anti-PD1 immunotherapy (pembrolizumab) according to current indications. Exclusion criteria were previous treatment for advanced disease or progression during neoadjuvant or adjuvant systemic treatment. Data from a cohort of healthy donors and of patients with NSCLC treated with anti-PD-1/PD-L1 after progression to first-line chemotherapy were also evaluated [16]. Ten ml of peripheral blood samples were obtained before the first cycle of immunotherapy, and after the first radiological control. PBMCs were isolated using Ficoll gradient as described elsewhere [16] and immune cell subpopulations were analyzed by flow cytometry. Participation of each patient in the study concluded when a radiological test confirmed response or progression, or if the patient withdrew consent or died. Tumor responses were evaluated according to RECIST 1.1 [17] and Immune-Related Response Criteria [18]. Progressive disease was confirmed by at least one sequential tumor assessment, except in the case of clear clinical deterioration.

### Flow cytometry

Surface flow cytometry analyses were performed as described elsewhere[19]. Blood samples (10 ml) were obtained from each patient, and immediately processed. PBMCs were isolated by FICOL gradients. PBMCs were washed and cells were stained with the indicated antibodies in a final volume of 50 μl for 10 min in ice. The following fluorochrome-conjugated antibodies were used at 1:50 dilutions unless otherwise stated: CD3-APC (ref 130-113-135, Miltenyi Biotech), CD4-APC-Cy7 (ref 130-113-251, Miltenyi Biotech), CD4-FITC (ref 130-114-531, Miltenyi Biotech), CD8-FITC (ref 35-0088-T500, Tonbo Biosciences), CD11b-PerCP-Cy5 (1:250) (ref 65-0112-U100, Tonbo Biosciences), CD14-PB (1:20) (ref 75-0149-T100, Tonbo Biosciences), CD27-PE (1:20) (ref 50-0279-T100, Tonbo Biosciences), CD28-PE-Cy7 (ref 302926, BioLegend), CD28-PerPC-Cy5 (1:20) (ref 302921, BioLegend), CD45RA-PB (ref 130-113-922, Miltenyi Biotech), CD56-PE-Cy7 (ref 130-113-870, Miltenyi Biotech), CD57-PB (ref 130-123-866, Miltenyi Biotech), CD62L-APC (1:20) (ref 304810, BioLegend), CD62L-PerCP-Cy5 (ref 304823, BioLegend), CD66b-APC-Cy7 (ref 130-120-146, Miltenyi Biotech), CD95-FITC (ref 130-124-261, Miltenyi Biotech), CD116-APC (ref 130-100-986, Miltenyi Biotech), CD119-PE (ref 130-125-874, Miltenyi Biotech), KLRG1-APC-Cy7 (ref 130-120-563, Miltenyi Biotech), LAG3-PE (ref 369306, BioLegend) and PD1-PE-Cy7 (ref 130-120-391, Miltenyi Biotech).

### Cell culture

Human lung adenocarcinoma A549 cells were a kind gift of Prof Rubén Pío, authenticated by his group, and were grown in standard conditions. They were confirmed to be mycoplasma-free by PCR. These cells were modified with a lentivector encoding a single-chain version of a membrane-bound anti-OKT3 antibody [20]. The lentivector expressed the single-chain antibody construct under the control of the SFFV promoter and puromycin resistance from the human ubiquitin promoter in a pDUAL lentivector construct [21]. The single-chain antibody construct contained the variable light and heavy OKT3 immunoglobulin sequences separated by a G-S linker fused to a human IgG1 constant region sequence followed by the PD-L1 transmembrane domain.

Cell growth and cytotoxicity were monitored using xCELLigence real-time cell analysis (RTCA ACEA Biosciences). Ten thousand A549-OKT3 cells resuspended in RPMI supplemented with 10% fetal bovine serum (FBS) were seeded in each well, adding plasma from patients or healthy donors at a 1:3 concentration when required by the experiment. After a 24-hour incubation, 5,000 T lymphocytes obtained from patients with NSCLC or healthy donors and stimulated with anti-CD3 and anti-CD28 were added. Each group had at least three repetitions, and each experiment was performed at least twice to confirm the results.

### Data collection and statistics

Immune cell subpopulations were quantified using FlowJo. The percentage of LDNs were quantified prior to therapy (baseline) and after the first radiological control. Data were recorded by H.A., A.B., M.Z. and M.G, and separately analyzed by H.A. and M.G.

Treatments were administered to the patients according to current indications. Progression-free survival (PFS) was defined as the time from the starting date of therapy to the date of disease progression or death from any cause, whichever occurred first. PFS was censored on the date of the last patient consultation when no signs of progressive disease were evident. PFS was represented by Kaplan-Meier Plots and log-rank tests were used to compare cohorts. Derived neutrophil-to-lymphocyte ratio (dNLR) and Lung Immune Prognostic Index (LIPI) score were calculated as described [22]. Receiver operating characteristics (ROC) analysis were performed with baseline LDNs numbers, NLR, dNLR and neutrophils and disease control at 6 months yes/no as a binary output. Overall survival (OS) was defined as the time from the start date of therapy to the date of death from any cause. OS was evaluated in the same way as PFS. Statistical tests were performed with GraphPad Prism 6 and SPSS statistical packages. Percentages of LDNs were not normally distributed, so the comparisons between groups were made using Mann-Whitney and Kruskal-Wallis tests.

### Proteomics

The Multiple Affinity Removal Spin Cartridge System (Agilent Technologies, Miami, USA) was used to remove the most abundant proteins according to the manufacturer’s instructions. Briefly, 6 μl of human plasma was diluted 16-fold with Buffer A and filtered through a 0.22-μm spin filter (1 min, 14,000 ×g). The non-bound protein fraction was collected and the column was washed twice with Buffer A and centrifuged (2.5 min, 100 ×g). Protein concentration was measured using Bradford assay kit (Biorad). MS/MS Library Generation and Quantitative Analysis were performed as previously described [23]. Briefly, 20 μg per sample were be used and the quantitative data obtained was analyzed using Perseus software [24] for statistical analysis and data visualization.

### Bioinformatics

Multiparametric flow cytometry data were represented in 2 dimensions using T-distributed Stochastic Neighbor Embedding (tSNE) algorithms [25]. Networks obtained from comparative proteomics of plasma from different cohorts were identified and represented using STRING (Search Tool for the Retrieval of Interacting Genes) software (http://stringdb.org/) [26].

## RESULTS

### 1. Study population

Thirty-one patients diagnosed with advanced NSCLC who received pembrolizumab as frontline therapy were recruited. All had PD-L1 tumor expression ≥ 50%. Baseline characteristics are depicted in Table 1. Overall response rate (ORR) was 42.9%, and disease control rate (DCR) was 64.3%. Median PFS and OS were 5.7 and 33.0 months, respectively. Fast progressive disease (fast-PD)/early death rate, defined as death at 12 weeks after starting, was 25.8%.

In another study cohort, 21 patients diagnosed with advanced NSCLC who received chemotherapy-immunotherapy combination (CT+IT) including pembrolizumab as first-line treatment were recruited. Overall, 61.9% had PD-L1 expression ≤ 1%, 14.3% had PD-L1 expression 1-4% and 23.8% had PD-L1 expression 5-49%. ORR was 47.4%, and DCR was 52.6%. Median PFS was 3.6 months, while median OS was not reached. The rate of fast-PD/early death was 9.5%.

### 2. Baseline low-density neutrophils (LDNs) and response to ICI monotherapy as frontline treatment in NSCLC

In the cohort of 31 patients diagnosed with advanced NSCLC who received pembrolizumab as frontline therapy, the overall immune cell composition in peripheral blood was characterized by high-dimensional flow cytometry and tSNE analyses in 3 consecutive patients with disease control longer than 6 months. These results were compared to those of 3 consecutive progressors. To avoid the confounding effect of chemotherapy, initially only patients receiving pembrolizumab monotherapy were evaluated.

Previous work by our group showed that relative percentages of highly differentiated CD4 T cells (CD4 T_HD_) in peripheral blood were a good predictive biomarker of response to second-line ICI monotherapies [16]. However, no correlation was observed in patients treated with frontline ICI therapy. Interestingly, a strong enrichment of low-density neutrophils (LDNs), a subpopulation with immunosuppressive properties identified as CD11b+ CD116+ CD66b+ CD3-CD14-, was found in progressors, while this population was apparently absent in responders (*Figure 1A*).

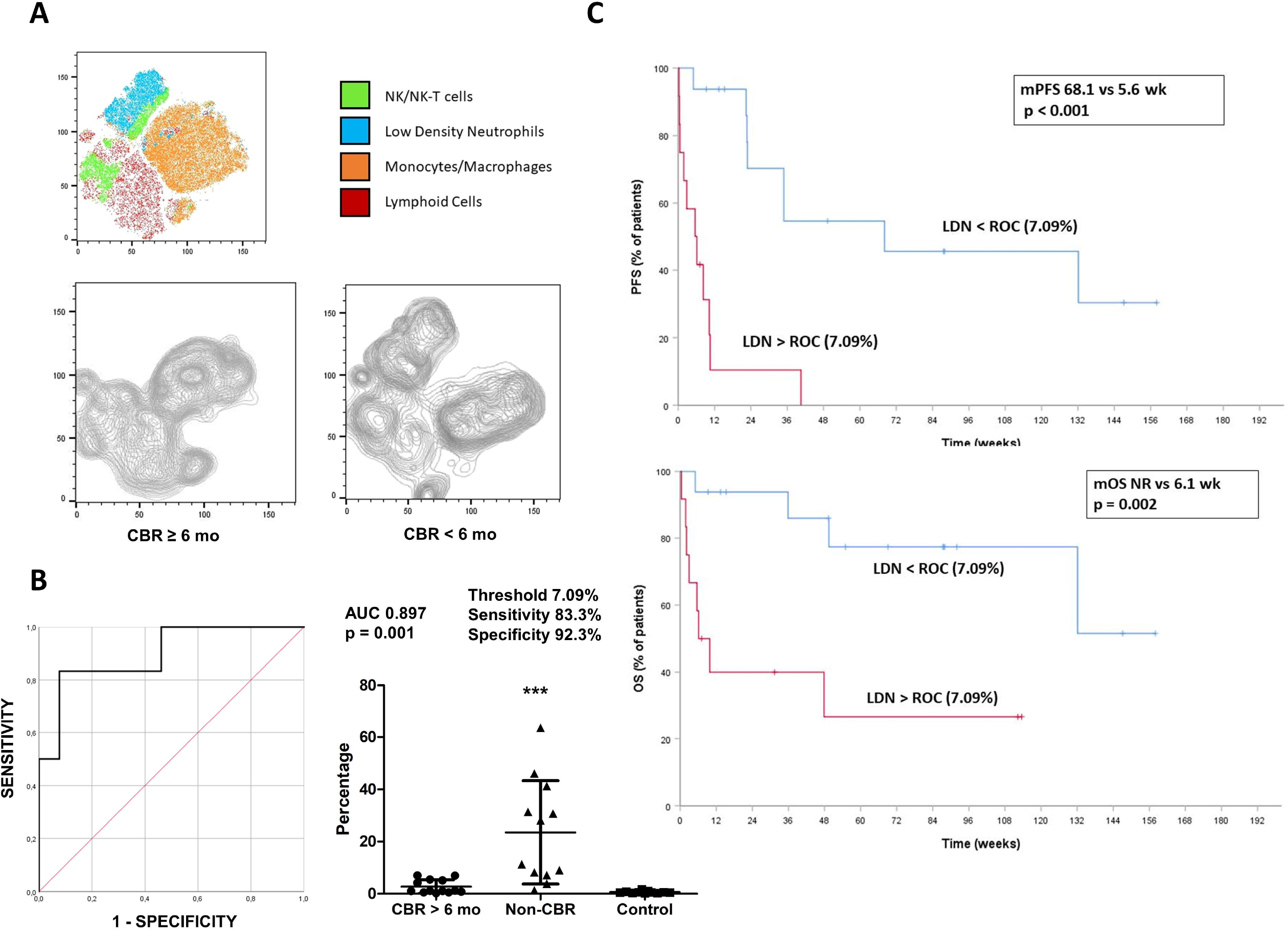

In the overall cohort of patients treated with ICI monotherapy, higher levels of LDNs were detected in progressors compared to responders or healthy donors (mean 23.5%, 2.7% and 0.7%, respectively; *p <0*.*0001*). To evaluate the value of baseline LDNs as a biomarker of response, ROC analyses were performed and an area under the curve (AUC) of 0.895 *(p = 0*.*001)* was calculated. ROC analyses established a LDN threshold of 7.09%, which identified patients who showed disease control of less than 6 months with a sensitivity of 83.3% and specificity of 92.3% *(Figure 1B)*. Patients with LDNs above this threshold presented an ORR of 0%, lower median progression free survival (mPFS) (6.1 weeks vs 68.1 weeks, *p <0*.*001*) and lower median OS (mOS) (6.1 weeks vs non reached, *p = 0*.*002*) (*Figure 1C*). Furthermore, the incidence of fast-PD/early death was significantly higher in patients with NSCLC and high LDNs (58.3% vs 6.3%, *p = 0*.*004)* (data not shown).

### 3. High LDN levels and resistance to first-line chemoimmunotherapy in patients with NSCLC

Twenty-one patients receiving chemoimmunotherapy combination (CT+IT) containing pembrolizumab as first-line treatment were recruited. In contrast to the results from first-line ICI monotherapy, baseline proportions of circulating LDNs did not predict response to CT+IT (AUC 0.471, *p = 0*.*845*), and levels did not differ between responders and non-responders (mean 9.8% and 12.8%, *p = 0*.*45*), suggesting a predictive and not prognostic value of LDNs (F*igure 2A*). Moreover, some patients with high baseline LDN levels (n = 9) responded to treatment and we observed in all of them a brisk decline in LDNs between the first and the second cycle. Significant proportions of LDNs were not longer observed at the time of the first radiological follow-up. This suggests that these cells play an active role in ICI resistance (*Figure 2B*).

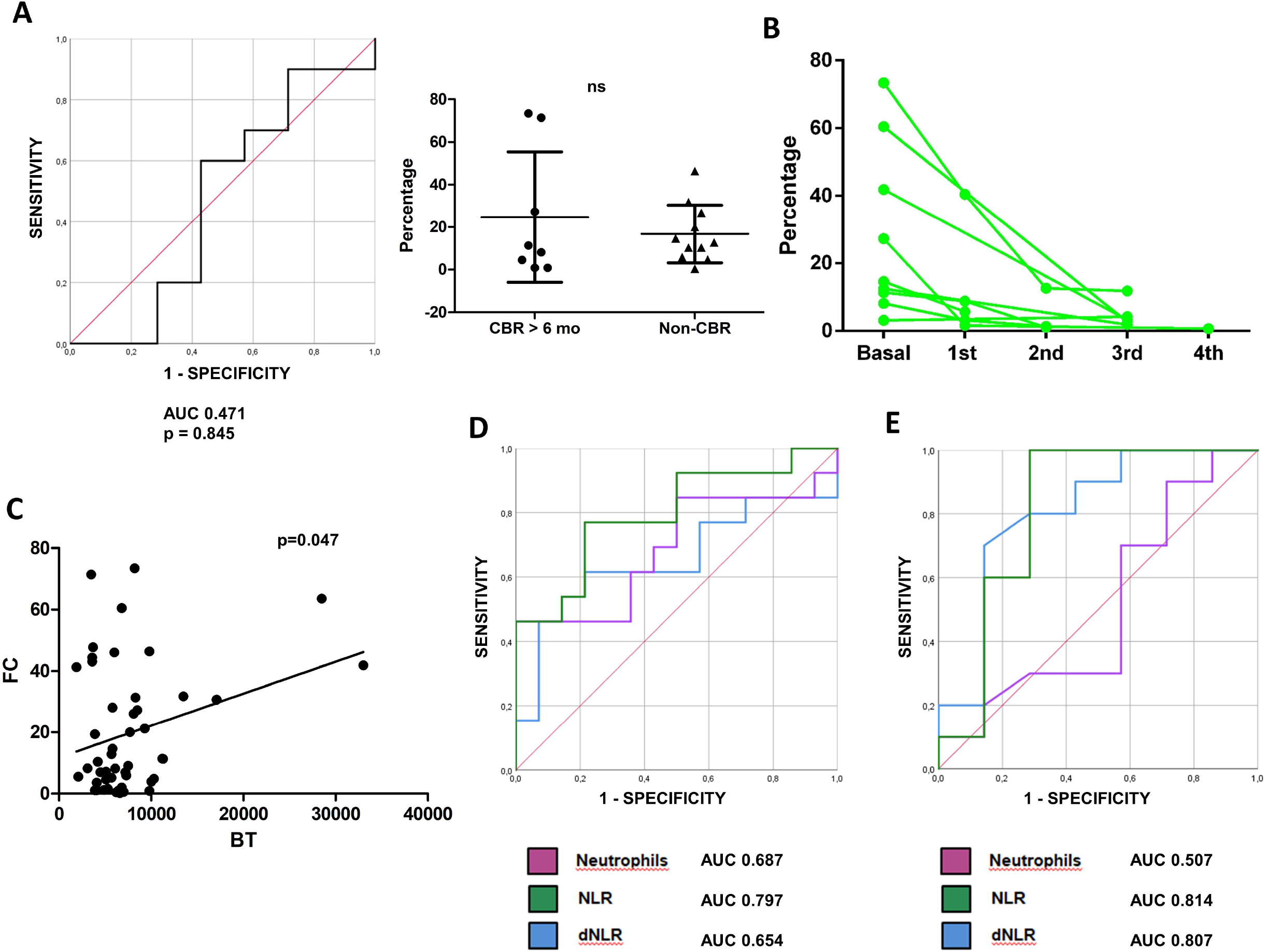

To confirm that the predictive value of LDNs was specific to patients treated with ICI monotherapy as first-line therapy, we retrospectively studied the flow cytometry stainings from a previous well-characterized cohort of patients with NSCLC treated with ICI monotherapy after progression to platinum-based chemotherapy [16]. No significant differences in LDNs were found between first-line and pretreated NSCLC (median 8.1% vs 4.4%, *p = 0*.*15*) (*data not shown*). Again, the mean proportion of LDNs was not higher in patients who progressed to treatment compared with responders (9.5% vs 19.1%, *p = 0*.*13*) (Figure 2C)

Previous works have demonstrated the predictive value of basaline neutrophils in peripheral blood tests from patients with NSCLC treated with ICI [22, 27-28]. To identify any relationship between neutrophils and LDNs, we studied the correlation between neutrophils from ordinary blood samples and LDNs quantified by flow cytometry. A statistically significant association was found by linear regression *(p = 0*.*047*), although the strength of the association was low. We observed that all patients with high levels of neutrophils presented a high proportion of LDNs (Figure 2C). However, 54% of patients with LDNs above the 7.08% threshold had no neutrophilia, suggesting that the expansion LDN expansion is independent of peripheral neutrophils.

The predictive value of peripheral neutrophils, neutrophil-to-lymphocyte ratio (NLR) and derived NLR (dNLR) was then studied. These variables were associated with progression to ICI monotherapy, although with lower sensitivity and specificity than LDNs (AUC 0.687, *p = 0*.*1*; AUC 0.797, *p = 0*.*01*; AUC 0.654, *p = 0*.*17*) (Figure 2D). Interestingly, NLR and dNLR were also associated with resistance to CT+IT (Figure 2E), and with OS in both the ICI monotherapy and the CT+IT cohort (not shown), indicating prognostic and not predictive value.

### 4. Soluble factors in plasma from patients with high LDN levels impair antitumor immune response ex vivo

To evaluate ex vivo T cell effector functions from the different cohorts of patients, we performed co-cultures of T cells with A549-SC3 cells, a human lung cancer cell line engineered by us to express membrane-bound anti-CD3 single-chain antibody to ensure tumor cell recognition by lymphocytes independently of antigen-specificity [16]. T cells from healthy donors were used as controls. T cells from patients with NSCLC regardless of their LDN status showed comparable cytotoxic activities to T cells from healthy donors (Figure 3A). These results showed that T cells from these patients are not dysfunctional per se.

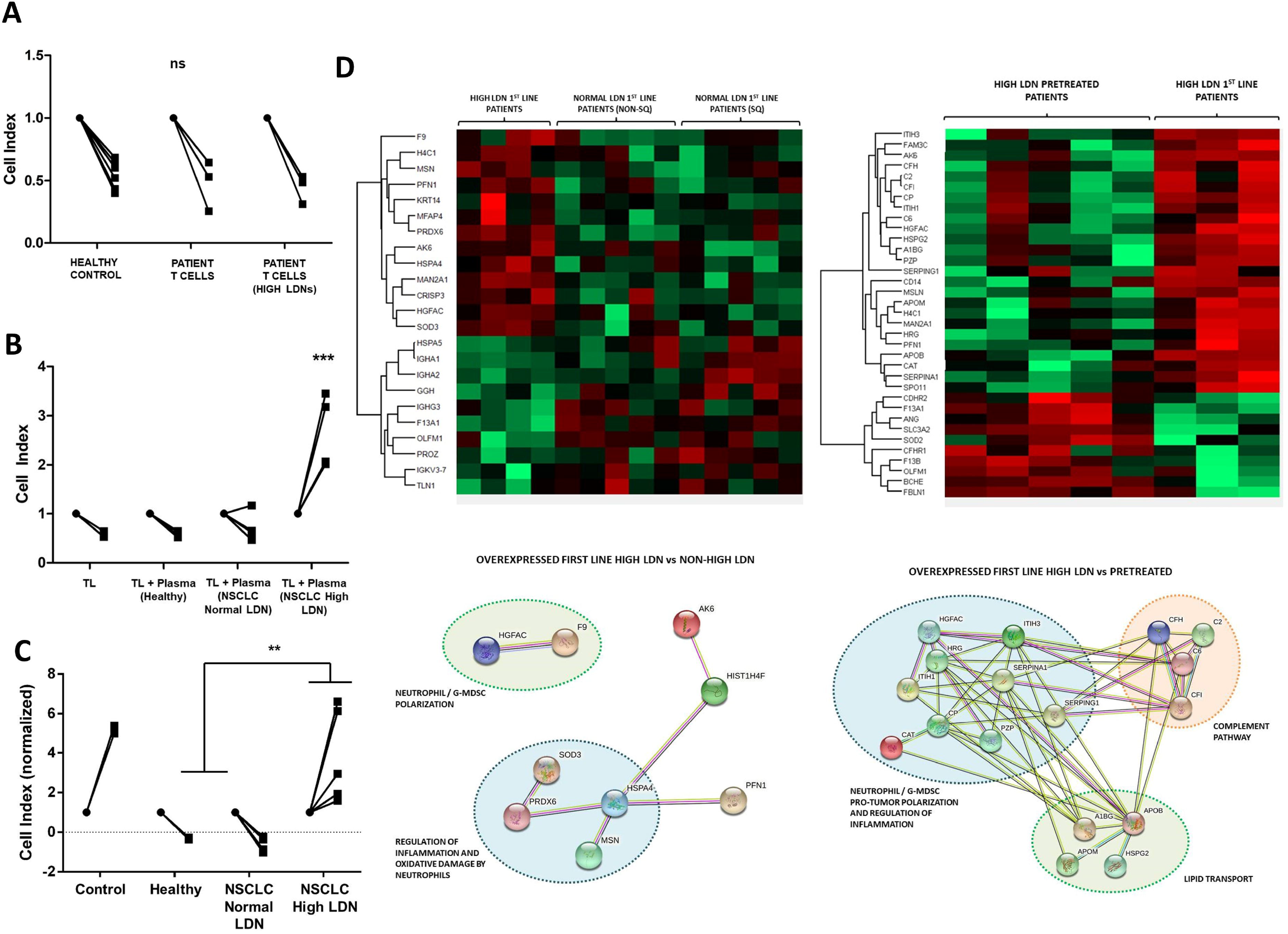

It had been previously demonstrated that soluble factors might mediate immune suppression caused by LDNs [29]. To find out if this was the case, co-cultures of A549-SC3 cells with T cells isolated from healthy donors were carried out in the presence or absence of plasma from healthy donors, from patients with NSCLC and normal LDNs proportions, and from patients with NSCLC and elevated LDNs. Interestingly, only plasma from patients with elevated LDNs levels abrogated T cell cytotoxicity over A549-SC3 cells (figure 3B). Furthermore, in absence of T cells, the plasma of these patients promoted cancer cell growth (Figure 3C).

Finally, to identify potential immunosuppressive candidates in plasma from patients with NSCLC and elevated LDNs, we performed quantitative proteomics in plasma. To this end, plasma from healthy donors, from untreated NSCLC patients with normal LDN proportions, from untreated patients with NSCLC and elevated LDNs, and from patients with NSCLC and elevated LDNs treated with ICI monotherapy after progression to platinum-based chemotherapy were used.

We found an enrichment in proteins related with neutrophil polarization and regulation of inflammation in untreated patients with NSCLC and elevated LDNs, confirming a potential role for LDNs in therapeutic resistance. Among these, hepatocyte-growth factor (HGF) activator (HGFAC), a serin-protease that activates HGF, was significantly elevated (*figure 3D*). HGF is an activator of the MET pathway, which has been associated with peripheral neutrophil expansion [30], immunosuppressive activity of these cells and lack of tumor T lymphocyte infiltration [29].

## DISCUSSION

The precise mechanisms underlying the anticancer immune response unleashed by immunotherapy are still not well characterized. Even though several resistance mechanisms have been described, their impact on the efficacy of the treatment in the different cancer subtypes is yet to be defined. Several papers exploring predictive biomarkers of response to ICI in NSCLC have been published up to date. However, the actual role that these biomarkers might play in the immune response has not been sufficiently detailed, probably due to the recruitment of heterogeneous cohorts of patients receiving different treatments in dissimilar clinical settings.

In agreement of other studies [31-33], we found a strong association between high baseline levels of circulating LDNs and resistance to ICI in untreated patients. LDN levels are higher in patients with advanced cancer compared with early-stage cancer patients or healthy controls [31-32]. Interestingly, LDNs were not associated with resistance to CT+IT, which indicates not only that circulating LDNs are a predictive biomarker of resistance to single-agent ICI as frontline therapy, but also suggests that chemotherapy combined with immunotherapy might deplete this cell population and allow an antitumor response. Neutrophil depletion, as well as IL-6 blockade, were associated with enhanced anti-PD1 immunotherapy efficacy in 2 lung cancer murine models [34-35]. Accordingly, we found that all patients with elevated LDNs who responded to CT+IT showed a brisk decline of LDNs after the first cycle, and this population was almost absent at the first radiological follow-up.

A recent paper reported higher rates of fast-PD/early death in patients with NSCLC and high dNLR before starting ICI monotherapy [36]. We found a numerically higher proportion of fast-PD/early death in the ICI monotherapy cohort than in the CT+IT cohort, and an association between fast-PD/early death and high LDNs in the ICI monotherapy cohort but not in the CT+IT cohort. This suggests that CT+IT treatment could prevent fast-PD/early death in patients with NSCLC and high LDNs receiving ICI monotherapy.

A translational study found that high blood neutrophil counts in melanoma patients refractory to immunotherapy were associated with high serum HGF levels. The authors demonstrated in a murine model that the inhibition of the HGF/c-MET axis impaired the recruitment of immunosuppressive neutrophils into tumors, thus allowing T cell tumor infiltration and enhancing the effect of immunotherapy [29]. Using ex vivo co-cultures we observed that plasma from patients with NSCLC and elevated LDN levels impaired the cytotoxic activities of T cells and promoted tumor cell proliferation. We used quantitative proteomics to compare the plasma of patients with elevated LDNs, and found an upregulation of proteins associated with the immunosuppressive role of LDNs, mainly bia the HGF/c-MET pathway.

Our findings show a specific association between LDNs and resistance to ICI monotherapy as frontline treatment in NSCLC. Patients with high LDN levels, regardless of high PD-L1 tumor expression, might benefit from CT+IT. The potential benefit of ICI combining with HGF/c-MET pathway inhibitors could be explored in further studies.

## Data Availability

All data produced in the present study are available upon reasonable request to the authors

## LIST OF ABBREVIATIONS

CT+IT: chemo-immunotherapy
dNLR: derived neutrophil-to-lymphocyte ratio
HGF: hepatocyte growth factor
HGFAC: HGF activator
ICI: immune checkpoint inhibitors
LDNs: low-density neutrophils
LIPI: Lung Immune Prognostic Index
NSCLC: non-small cell lung cancer
OS: overall survival
PFS: progression-free survival
ROC: receiver operating characteristics
STRING: Search Tool for the Retrieval of Interacting Genes
tSNE: T-distributed Stochastic Neighbor Embedding

## KEY MESSAGES

1. High baseline low-density neutrophils identify a group of patients with non-small cell lung cancer intrinsically refractory to single anti-PD1 monotherapy. These patients can be effectively treated with chemoimmunotherapy combination.
2. Depletion of low-density neutrophils is observed in patients who responded to chemoimmunotherapy.
3. This phenomenon is caused by soluble compounds present in patients’ plasma. Comparative quantitative proteomics indicate an upregulation of the HGF/c-MET pathway in patients with elevated low-density neutrophils.

## ACKNOWLEDGMENTS

We sincerely thank the patients and families who generously agreed to take part in this study. We also thank the nursing staff of the Medical Oncology Day Care at University Hospital of Navarre for their willful collaboration, and the Blood and Tissue Bank of Navarre, Health Department of Navarre, Spain. We are thankful of the support provided by PUBLIbeCA SEOM 2020 from the Spanish Society of Medical Oncology (SEOM) for the writing of the article.

