## Supplementary material for "Circulating Low Density Neutrophils Are Associated with Resistance to First-Line Anti-PD1/PDL1 Immunotherapy in Non-Small Cell Lung Cancer": Figures

Figure 1. A: Top, tSNE graph of myeloid subpopulations represented by different colors. Lower left, tSNE graph of 3 responders to immunecheckpoint inhibitors (ICI) monotherapy. Lower right, tSNE graph representative of 3 non-responders to ICI monotherapy. B: Left, ROC analysis of baseline low-density neutrophils (LDNs) as a function of clinical benefit rate (CBR) < 6 months. Right, baseline levels of LDN in patients receiving ICI monotherapy with CBR longer than 6 months compared with patients with CBR less than 6 months and with healthy donors. C: Top, progression free survival stratified by the presence of baseline LDNs above the ROC threshold (7.09%). Below, overall survival stratified by the presence of baseline LDN above the ROC threshold.

Figure 2. A: Left, ROC analysis of baseline LDNs as a function of CBR < 6 months in patients receiving chemoimmunotherapy (CT+IT). Right, baseline levels of LDN in patients receiving CT+IT with CBR for more than 6 months compared to patients with CBR less than 6 months. B: Monitoring of patients with LDNs above the threshold who responded to CT+IT. C: Scatter plot representing the association between baseline LDNs and baseline neutrophils in blood test in the whole group of patients. D: ROC analysis of baseline neutrophils, neutrophil-to-lymphocyte ratio (NLR) and derived neutrophil-to-lymphocyte ratio (dNLR) as a function of CBR < 6 months in patients receiving ICI monotherapy. E: ROC analysis of baseline neutrophils, NLR and dNLR as a function of CBR < 6 months in patients receiving CT+IT.

Figure 3. A: Cell index of A549-OKT3 cells before and after the addition of T lymphocytes from the cohorts indicated below. B: Cell index before and after the addition of T lymphocytes from a healthy donor and plasma from the cohorts indicated below. C: Cell index before and after the addition of plasma from the cohorts indicated below. D: Upper left, heat-map comparing the complete identified proteome from the plasma of untreated NSCLC patients with high baseline LDN levels, of untreated squamous NSCLC patients with normal LDN levels and of untreated non-squamous NSCLC patients with normal LDN levels. Top right, heatmap comparing the complete identified proteome from the plasma of untreated NSCLC patients with high baseline LDN levels, with that of NSCLC patients with high LDN levels who progressed to platinum-based chemotherapy. Lower left, functional interactomes with significantly upregulated proteins in untreated NSCLC patients with high baseline LDN levels compared to untreated NSCLC patients with normal baseline LDN levels. Lower right, functional interactomes with significantly upregulated proteins in untreated NSCLC patients with high baseline LDN levels compared to NSCLC patients with normal baseline LDN levels who progressed on platinum-based chemotherapy.
